# Beyond Hesitancy: Assessing the vaccination gap among Children and Livestock in the Maasai community of Kajiado, Kenya

**DOI:** 10.64898/2026.06.30.26356974

**Authors:** Peris Kung’u, Victor Mbao, Samuel Oji Oti

## Abstract

Despite established national immunization programmes for children and livestock, pastoral communities in Kenya remain chronically underserved, with low vaccination coverage attributed mainly due to seasonal mobility, vast terrain, and limited health infrastructure. Even less understood is whether these assumptions hold across all pastoral contexts, and how livelihood practices such as hiring herders during drought seasons may intersect with vaccine access. This study examined factors associated with child and livestock vaccination among Maasai communities in Kajiado Central Subcounty, Kenya, using a One Health lens.

We employed a mixed-methods design across three wards (Dalalekutuk, Ildamat, and Purko). Quantitative data were collected through semi-structured household surveys (n=180), with bivariate analysis examining the associations between the vaccine hesitant group and the vaccine accepting group in children and livestock. Qualitative data were gathered through two gendered focus group discussions (FGDs, n=31) and seven key informant interviews (KIIs). Inductive thematic analysis was interpreted through the COM-B framework, and findings were integrated using convergent triangulation.

Child immunization coverage averaged 90% (range 87–98%), which is higher than typically reported for pastoral populations. In contrast, livestock vaccination coverage averaged 53% (range 5–87%) despite comparable willingness to vaccinate in both children (97%) and livestock (93%). Vaccine hesitancy co-occurred across children and livestock within the same households (OR 36.7, 95% CI 5.9 - 227.5). Eighty-eight percent of households hired herders to migrate with livestock during the cool-dry season (June–September), suggesting a shift toward sedentarization. Qualitatively, supply chain failures including vaccine production monopoly, counterfeit vaccines, stockouts, and understaffing were identified as key contributors to low livestock vaccination coverage.

Closing the livestock vaccination gap requires supply-chain reforms such as breaking the KEVEVAPI monopoly, strengthening the VMD regulatory framework, and securing transport budgets to avoid stockouts. The relationship between hiring herders and vaccine access warrants further investigation as a potential structural enabler towards strengthening pastoral health programming.

**AI use declaration:** AI (Chat GPT) was used to improve flow and grammar of this manuscript. All scientific content, data analysis, and interpretation are the original work of the authors.

## Introduction

Vaccination for children and livestock remains one of the most impactful and cost-effective public health interventions globally, yet coverage continues to be uneven across populations(1,2). In Kenya, the national child immunization programme delivers a comprehensive routine schedule including: Bacillus Calmette–Guérin (BCG), Oral Polio Vaccine (OPV), Inactivated Polio Vaccine (IPV), Pentavalent (Diphtheria, Pertussis, Tetanus, Hepatitis B, Haemophilus influenzae), Pneumococcal Conjugate Vaccine (PCV10), Rotavirus, and Measles-Rubella vaccines through public and private health facilities(3). In contrast, livestock vaccination in Kenya is managed at the sub-national (county) level, with the Ministry of Agriculture, Livestock and Fisheries (MALF) providing routine vaccines against prevalent diseases in specific counties. For instance, in Kajiado County, eleven livestock diseases have been identified and ranked by importance, of which the top three are vaccine-preventable: Contagious Caprine Pleuropneumonia (CCPP), Lumpy Skin Disease (LSD), and Foot and Mouth disease (FMD)(4). Yet these programmes are largely designed for sedentary populations, and their reach has remained uneven especially among the pastoral communities.

Their dispersed settlement patterns, seasonal mobility, and limited engagement with formal health systems create persistent barriers to both human and animal health service access(5,6). Child immunization coverage among pastoralists has consistently been shown to fall below national averages, driven by the mobility of women and children, prevailing cultural norms, and constrained access to services(7–9). On the livestock side, research in the Southern Maasai rangelands identified institutional failures including inadequate government service provision, long distances to vaccine suppliers (e.g agrovet), weak institutional linkages, and absent cold-chain infrastructure as the primary barriers to accessing vaccines(10). Significantly, pastoralists in these settings ranked vaccines among their most critical inputs and expressed willingness to pay for accessible, integrated services, pointing to a demand that supply systems have failed to meet(10).

What remains insufficiently examined is how livelihood practices (particularly drought-driven livestock management) interact with vaccine access. The question of whether a One Health framework can reveal shared structural determinants of vaccine acceptance across humans and animals remains a significant evidence gap in pastoral vaccination programming(2,11). Additionally, separating supply-side from demand-side contributors is equally crucial, as each calls for a different programmatic response(12).

A One Health approach offers a combined framework to address these gaps, integrating human and animal health systems in a way that reflects the realities of agropastoral communities(2,13). Therefore, this study aimed to identify the factors associated with child and livestock vaccination uptake among Maasai communities in Kajiado Central Subcounty using a mixed-methods design. Specifically, it examined vaccination coverage levels, identified structural and behavioral barriers and facilitators of uptake, and explored how social factors shape vaccine acceptance across both humans and animals. By integrating quantitative and qualitative evidence, the study sought to generate context-specific insights to inform tailored vaccination strategies for pastoralist communities in Kenya.

## Methodology

### Study Site

The study was conducted in Dalalekutuk, Ildamat, and Purko wards of Kajiado Central Subcounty in Kajiado county, located in the southern part of Kenya. The county hosts a large population of pastoralists from the Maasai community. Figure 1 shows a map of the study area.

**Figure 1:**
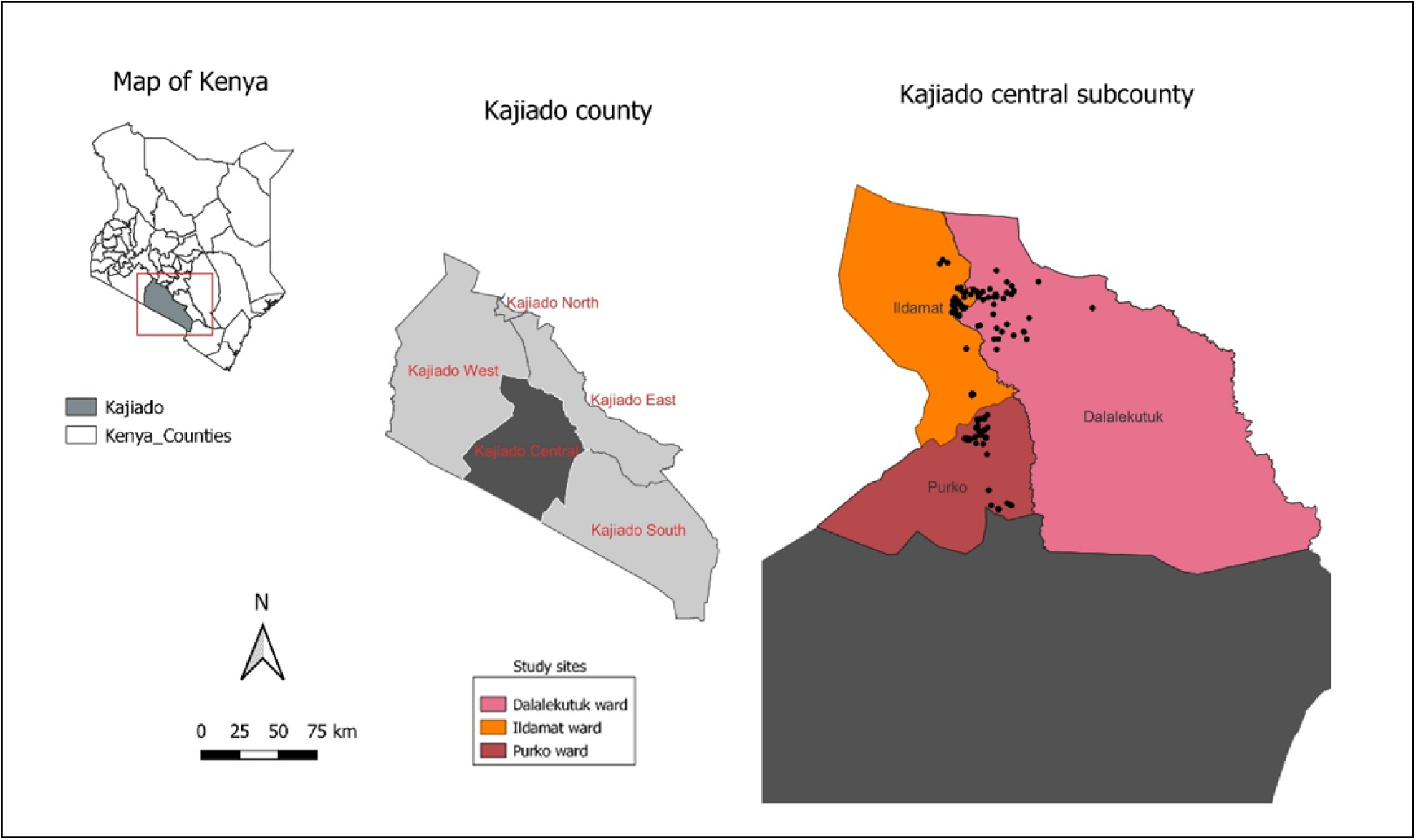
A map illustrating the study area, Kajiado Central Subcounty

### Study Design

The study employed a mixed-method approach comprising both quantitative and qualitative data collected. Quantitative data was collected through cross sectional household surveys while qualitative data was gathered through focus group discussions (FGDs) and key informant interviews (KIIs).

### Sampling

Eligible households were identified using non-discriminatory exponential snowball sampling, a form of purposive chain-referral sampling suited for hard-to-reach pastoralist populations leveraging existing social networks. These eligible households were recruited from 15^th^ January to 29^th^ February 2024. With the assistance of a local *nyumba kumi* (village) leader, an initial eligible household (defined as having at least one child under five years and owning at least one livestock) was initially identified. That household then referred the enumerator to additional eligible households, and each newly referred household provided 1 or >1 referrals until the target sample size was reached.

The target of 180 households was guided by practical constraints including the two-month field period and available budget. No prior power calculation was performed, as the study was exploratory, the prevalence of vaccine hesitancy in this population was not known. The primary objectives were descriptive (estimating coverage levels) and bivariate (comparing proportions across subgroups). Nevertheless, the sample of 180 households was considered sufficient to detect moderate differences in key comparisons such as vaccine hesitant by vaccine accepting households and to capture variation across the three study wards.

### Data collection

For the household survey, a semi–structured questionnaire (English and Kiswahili) was administered by six trained enumerators to 180 households in Kajiado Central Subcounty. The questionnaire collected information on sociodemographic, socioeconomic, health–seeking behaviour, and knowledge, attitudes and practices regarding child immunization and livestock vaccination.

Qualitative data were collected using two focus group discussions (FGDs) and seven key informant interviews (KIIs). Two FGDs were conducted: one with female participants (n=17) and one with male participants (n=14), distributed across age groups: 20–30 years (n=15), 31–40 years (n=8), 41–50 years (n=3), and 50+ years (n=5). Each FGD lasted 1– 2 hours and was facilitated by two trained moderators using a guide (English and Kiswahili) that covered cultural beliefs, barriers, facilitators, and recommendations for child and livestock vaccination.

KII guides were customized to the informant category. For technical experts (Subcounty Expanded Programme on Immunization nurse, Subcounty public health officer, Subcounty veterinary officer in charge, and veterinary officer in charge of stores), questions focused on vaccine supply chains, cold chain logistics, and coverage trends. For community and religious leaders (a farmer, a bishop, and a sheikh), questions explored local perceptions, barriers, facilitators, and recommendations.

All FGDs and KIIs were audio–recorded, transcribed verbatim, and translated into English where needed.

### Data Analysis

Quantitative data were analyzed using R software (version 4.2.2). Continuous variables were summarized as means or medians, and their distributions were assessed graphically. Categorical variables were summarized using percentages and frequencies. Bivariate associations between categorical variables were examined using chi-square tests (or Fisher’s exact test when any expected cell count was <10). Variables with a bivariate p-value <0.05 were considered for inclusion in multivariable logistic regression models.

However, due to the small number of hesitant households in both child (n=6) and livestock (n=13) hesitancy outcomes, multivariable logistic regression was not feasible for either outcome. For both, unadjusted odds ratios were calculated from 2×2 contingency tables and findings were reported descriptively. Complete separation on multiple variables further precluded model estimation; where this occurred, only p-values from bivariate tests were reported.

An inductive thematic analysis approach was used for the qualitative analysis (14). Transcripts were loaded into Taguette for coding, with codes being grouped into themes and further organized using Microsoft Excel. This analysis focused on recurrent patterns in vaccine acceptance, barriers to access, gendered decision making, and community perceptions of child and livestock vaccination. Following the inductive coding, themes were deductively interpreted through the COM-B framework (Capability, Opportunity, Motivation-Behaviour) to examine how behavioural and structural determinants shape vaccine hesitancy or acceptance in children and livestock.

Integration of quantitative and qualitative data was attained through convergent triangulation (Figure 2)(15). We compared quantitative patterns (p-values, proportions, odds ratios) with the qualitative themes (barriers, facilitators) to determine points of convergence and divergence. A joint display table summarizing key integrated findings was generated.

**Figure 2:**
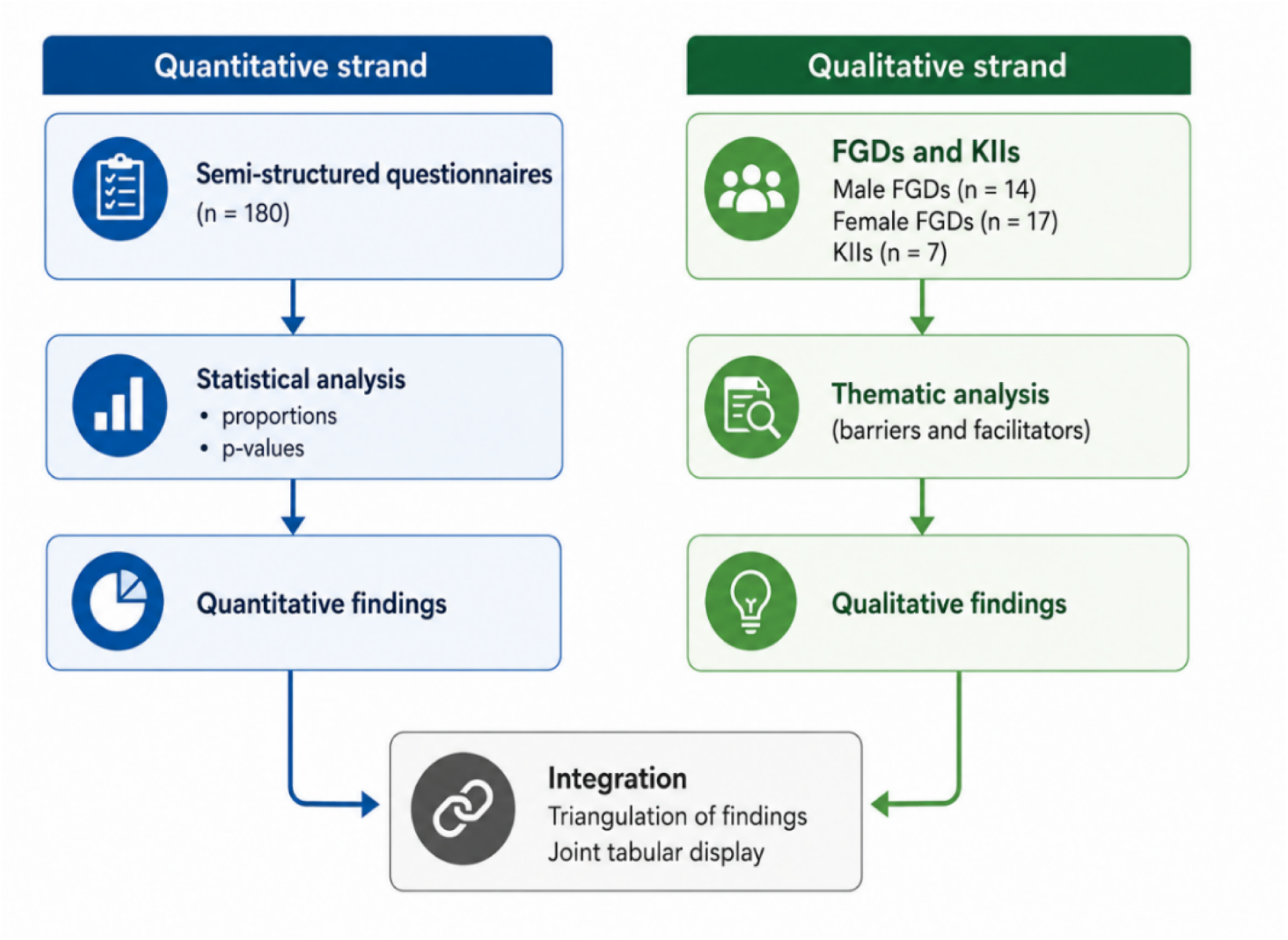
Schematic representation of the convergent triangulation approach used to integrate quantitative survey data with qualitative FGD and KII findings.

## Ethical Considerations

Ethical approval was obtained from the Amref Ethics and Scientific Review Committee (ESRC P1497/2023). A preliminary visit was conducted to obtain Kajiado County administrative permission and to pilot the questionnaire and FGD/KII guides. The tools were tested with four participants in Ildamat ward and adjusted accordingly. All participants provided a written informed consent (thumbprints were obtained from non-literate participants). Confidentiality was maintained through anonymization of the data using unique codes, with all the audio recordings and transcripts stored on password protected files.

## Results Quantitative results

### Characteristics of participants

Out of 182 participants interviewed from Kajiado Central subcounty, 180 completed the questionnaire. Each household had between 3 and 21 residents, with the most common sizes being 6 (17%, n=31), 5 (16%, n=29), and 7 (12%, n=21) individuals.

Table 1 represents the characteristics of the 180 participants stratified by gender. Sixty three percent (n=113) of the participants were females. A notable education gap (by gender) was observed with 38% (n= 43) females having no formal education compared to 28% (n= 19) males and only 4.4% (n= 5) females compared to 22% (n=15) attaining tertiary education.

**Table 1.**
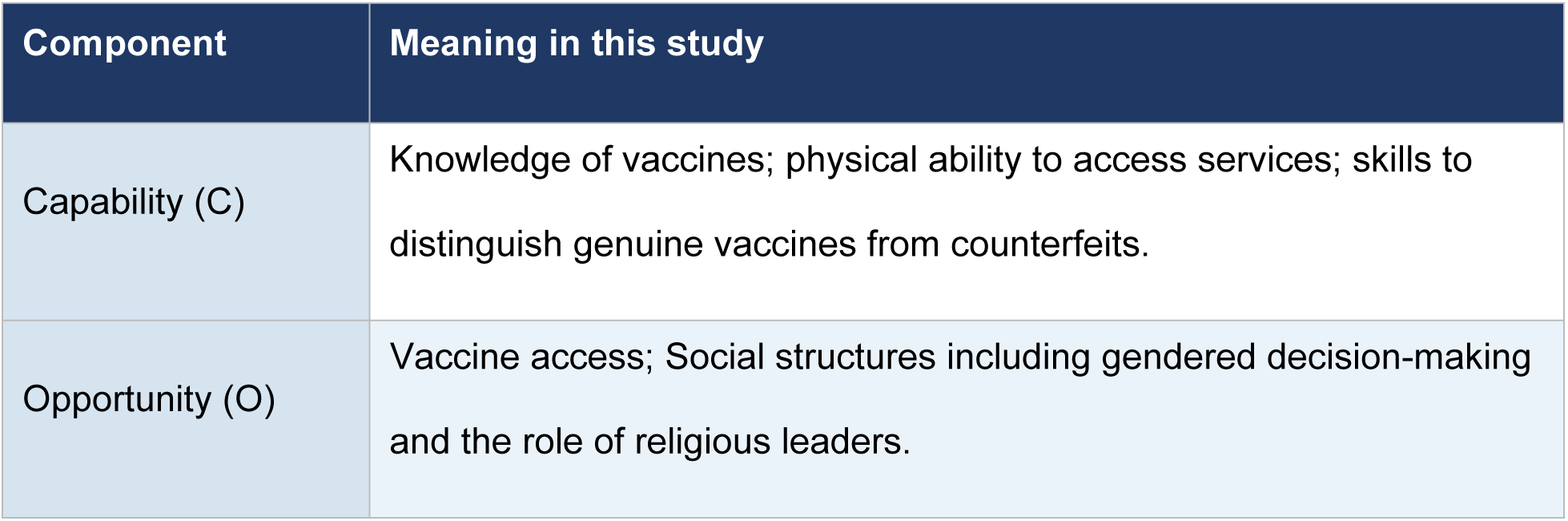
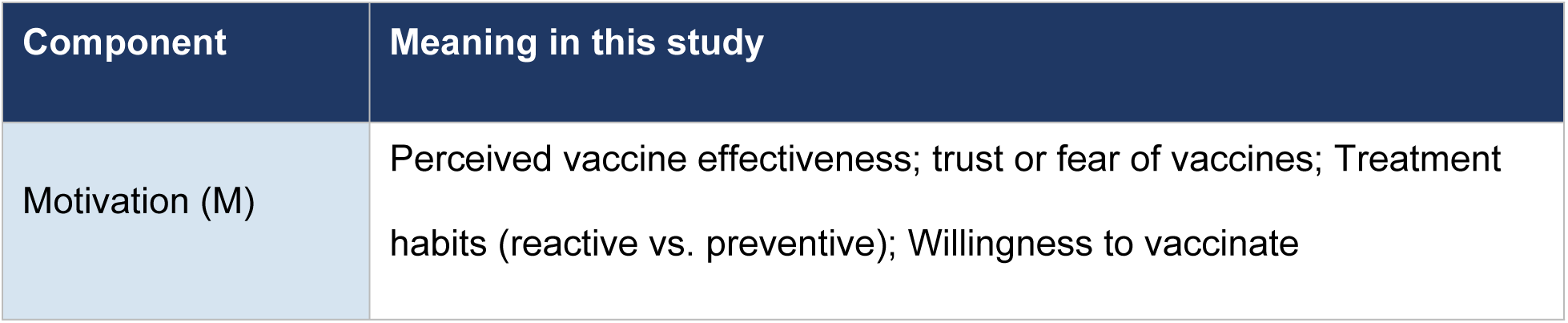
COM-B framework as operationalized in this study.

| Component | Meaning in this study |
| --- | --- |
| Capability (C) | Knowledge of vaccines; physical ability to access services; skills to distinguish genuine vaccines from counterfeits. |
| Opportunity (O) | Vaccine access; Social structures including gendered decision-making and the role of religious leaders. |
| Motivation (M) | Perceived vaccine effectiveness; trust or fear of vaccines; Treatment habits (reactive vs. preventive); Willingness to vaccinate |

**Table 2:** Characteristics of the 180 participants by gender.

| Characteristic | Female (n=113) | Male (n=67) | Total (N=180) |
| --- | --- | --- | --- |
| **Ward** |  |  |  |
| Dalalekutuk | 45 (39%) | 17 (25%) | 62 (34%) |
| Ildamat | 41 (36%) | 18 (27%) | 59 (33%) |
| Purko | 28 (25%) | 32 (48%) | 60 (33%) |
| **Age group** |  |  |  |
| 18–34 years | 71 (63%) | 30 (45%) | 101 (56%) |
| 35–54 years | 40 (35%) | 37 (55%) | 77 (43%) |
| 55 and above | 2 (1.8%) | 0 (0%) | 2 (1%) |
| **Education** |  |  |  |
| No formal education | 43 (38%) | 19 (28%) | 62 (34%) |
| Primary school | 32 (28%) | 11 (16%) | 43 (24%) |
| Secondary school | 33 (29%) | 22 (33%) | 55 (31%) |
| Tertiary/Higher | 5 (4.4%) | 15 (22%) | 20 (11%) |
| **Marital status** |  |  |  |
| Married | 97 (86%) | 66 (99%) | 163 (91%) |
| Single guardian | 16 (14%) | 1 (1.5%) | 17 (9%) |
| **Main income source** |  |  |  |
| Livestock farmer | 65 (58%) | 36 (54%) | 101 (56%) |
| Employment | 13 (12%) | 15 (22%) | 28 (16%) |
| Skilled labour | 17 (15%) | 11 (16%) | 28 (16%) |
| Herder | 9 (8.0%) | 3 (4.5%) | 12 (7%) |
| Crop farmer | 8 (7.1%) | 1 (1.5%) | 9 (5%) |
| **Season of migration** |  |  |  |
| June–September<br>(Cool/dry) | 82 (73%) | 42 (63%) | 124 (69%) |
| January–February<br>(Hot/dry) | 30 (27%) | 23 (34%) | 53 (29%) |

Fifty-six percent (n=101) aged between 18-34 years, with 91% (n= 163) being married. The median monthly income among participants was Ksh 15,000 (range Ksh 50–250,000). Livestock farming was the main source of income for 56% (n=101) of the participants. Sixty nine percent (n= 124) of the participants migrated seasonally primarily during the cool and dry season (June-September).

### Child Immunization

Fifty-seven percent (n=102) of children born were male. Ninety-one percent (n=164) of the participants presented child immunization cards. Table 3 presents the coverage estimates based on card documentation, supplemented by caregiver recall in the absence of the cards. Among eligible children that received each vaccine, coverage was 98% for BCG, 92% for OPV0 while for the first-year vaccines including: OPV1-3, Pentavalent 1-3, PCV 1-3, Rotavirus, coverage ranged from 87% to 92%. The average immunization coverage is 90%.

**Table 3.**
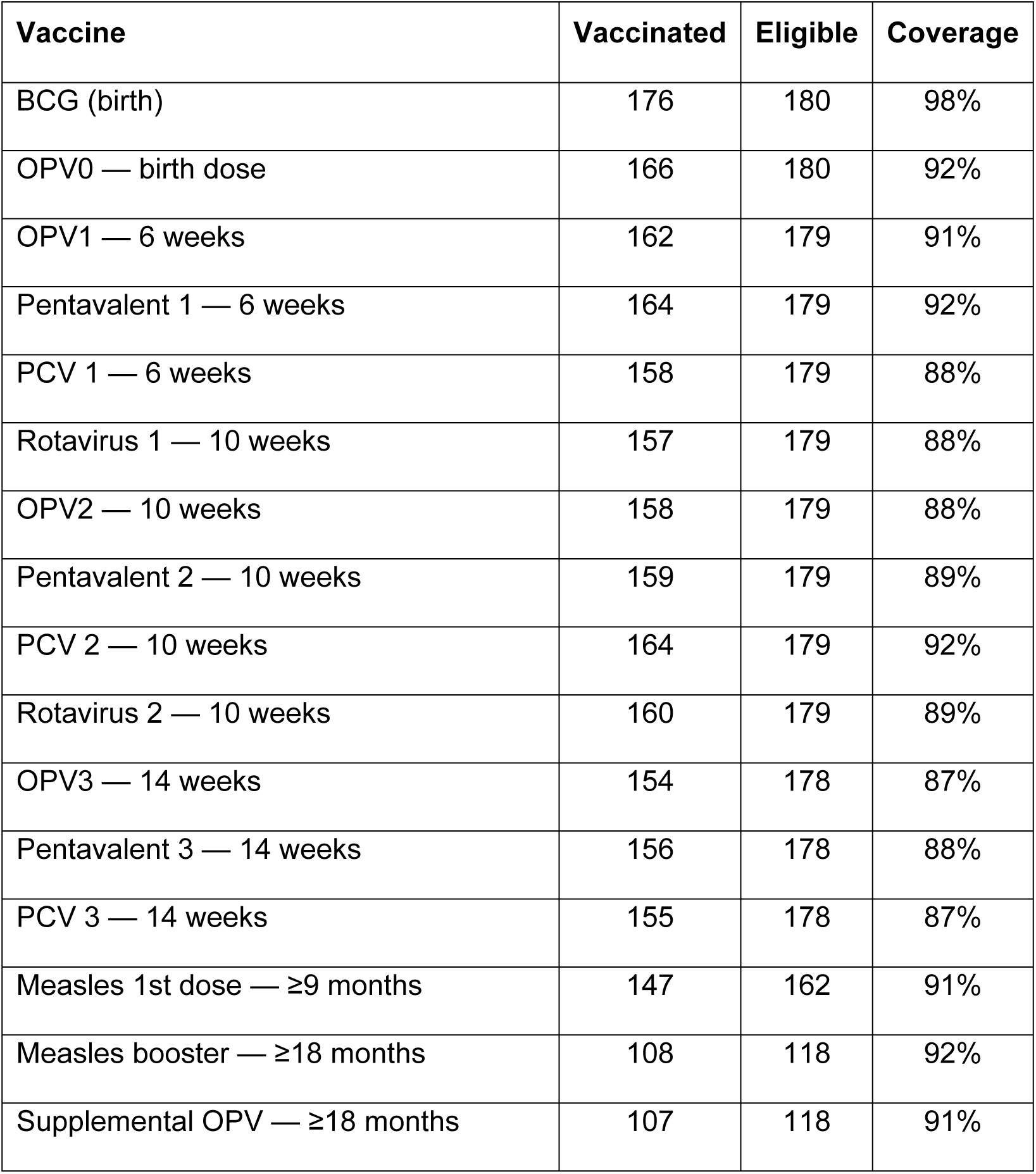
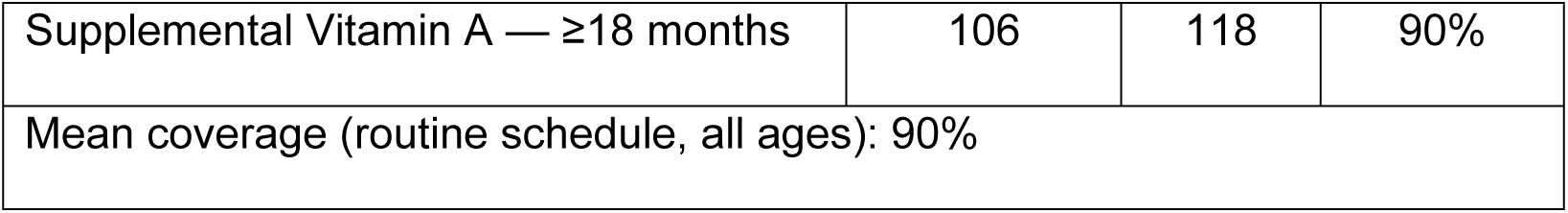
Vaccine-specific coverage among children under five (N= 180 HH), according to the Kenya National Immunization Schedule.

The dominant source of child immunization information was the radio stations cited by 91% (n=164) participants. Sixty seven percent (n=120) of the participants travelled to health facilities via motorcycle at a median cost of Ksh 150 (IQR 50-300), while 47% (n=84) walked, 11% (n=20) used a private vehicle and only 8% (n=14) used a bus. Ninety seven percent (n=174) of the participants were willing to immunize their children.

### Child vaccine hesitancy

Six households (3.3%) reported child vaccine hesitancy. Four of the six (67%) also reported livestock vaccine hesitancy, indicating co-hesitancy. Table 4 presents the bivariate comparison of child-hesitant and child-accepting households. Livestock vaccine hesitancy (p<0.001), non-receipt of the November–December 2023 polio vaccine (p=0.005), and Travel cost (p= 0.036) were significantly associated with child hesitancy.

**Table 4.** Bivariate analysis: child hesitant vs child accepting households (N=180).

| Characteristic | Hesitant (n=6) | Accepting (n=174) | P-value |
| --- | --- | --- | --- |
| **One Health: Co-Hesitancy** |  |  |  |
| Livestock Vaccine Hesitancy (Yes) | 4 (67%) | 9 (5%) | **<0.001** |
| **Clinical History** |  |  |  |
| Polio vaccine received Nov–Dec 2023 | 4 (67%) | 172 (99%) | **0.005** |

| Access & Transport |  |  |  |
| --- | --- | --- | --- |
| Travel Time ≥1 Hour to Facility | 4 (67%) | 75 (43%) | 0.407 |
| Travel Cost (Ksh) — Median (IQR) | 175 (100–225) | 150 (50–300) | **0.036** |

*Fisher's exact test throughout. Bold = $p < 0.05$*

Due to complete separation on multiple variables and an events per variable (EPV) ratio of 6, multivariable logistic regression was not estimated; findings are therefore reported descriptively and through bivariate analysis only. We conducted a 2×2 table for co–hesitancy (child hesitant vs livestock hesitant) which yielded an unadjusted odds ratio of 36.7 (95% CI: 5.9–227.5, p<0.001). While this indicates a strong association between child and livestock vaccine hesitancy, the wide confidence interval reflects small cell counts and the estimates should be interpreted with caution.

### Livestock vaccination

Most participants (82%, n=147) kept sheep, goats, and cattle with average holdings of 19 sheep, 16 goats, and 10 cattle per household. Livestock vaccination coverage varied considerably by disease, with the top three diseases (LSD 87% n=140, FMD 82% n=134, CCPP 69% n=111) achieving the highest coverage, followed by BQ 65% (n=104), PPR 51% (n=92), SNG pox 47% (n=85). While Newcastle disease and enterotoxaemia had coverage below 10% (Table 5). The mean livestock vaccination coverage was 53%, which was 37% lower than child immunization (90%).

**Table 5.** Livestock vaccination coverage and disease priority ranking (N=180 HH)

| Disease | Species | Eligible | Vaccinated | Not Vaccinated | Coverage |
| --- | --- | --- | --- | --- | --- |
| LSD | Cattle | 161 | 140 | 21 | 87% |
| FMD | Cattle | 161 | 134 | 27 | 83% |
| CCPP | Cattle | 161 | 111 | 50 | 69% |
| BQ | Cattle | 161 | 104 | 57 | 65% |
| PPR | Sheep/Goats | 175 | 90 | 85 | 51% |
| SNG pox | Sheep/Goats | 175 | 82 | 93 | 47% |
| Newcastle Disease | Poultry | 91 | 13 | 78 | 14% |
| Enterotoxemia | Sheep/Goats | 175 | 8 | 167 | 5% |
| Mean livestock vaccination coverage = 53% |  |  |  |  |  |

The primary source of livestock vaccination information was word of mouth through village elders and the church (71%, n=128), contrasting with radio dominance for child immunization (91%). Eighty-eight percent (n=159) of participants hired herders to move with their livestock during the drought season. Willingness to vaccinate livestock was high at 93% (n=167).

Thirteen households (7.2%) reported livestock vaccine hesitancy. Of these, 62% (8/13) had never received a visit from animal health workers, compared to 34% (57/167) of accepting households. Thirty-one percent (4/13) also reported child vaccine hesitancy. Table 6 presents bivariate comparisons for livestock hesitancy.

**Table 6.** Bivariate analysis: livestock vaccine hesitant vs livestock vaccine accepting households (N=180).

| Characteristic | Hesitant (n=13) | Accepting (n=167) | P-value |
| --- | --- | --- | --- |
| **One Health: co-hesitancy** |  |  |  |
| Child vaccine hesitancy (Yes) | 4 (31%) | 2 (1%) | **<0.001** |
| **Animal Health Service Providers (AHSP) visits** |  |  |  |
| AHSP visits: Never | 8 (62%) | 57 (34%) | 0.095* |
| AHSP visits: Every 6 months | 0 (0%) | 57 (34%) |  |
| **Distance to livestock vaccination point** |  |  |  |
| Below 5 km | 11 (85%) | 97 (58%) | 0.267* |
| Years rearing animals — median (IQR) | 15 (10–18) | 15 (10–20) | 0.411 |

*Fisher's exact test or chi-square as appropriate. \*Overall p-value. Bold = $p < 0.05$ .*

Due to the small number of events (n=13) and complete separation on several variables, multivariable logistic regression was not feasible; findings were therefore reported descriptively and through unadjusted analysis only. A 2×2 contingency table examined the association between child and livestock vaccine hesitancy. Child vaccine hesitancy was strongly associated with livestock vaccine hesitancy, yielding an unadjusted odds ratio of 36.67 (95% CI: 5.91–227.47, p<0.001). Households that had never received an animal health worker (AHSP) visit were more common in the hesitant group (62%, 8/13) than in the accepting group (34%, 57/167) (p=0.095).

## Qualitative results

Five themes emerged after conducting the inductive analysis on the FGD and KII data. Verbatim quotations were labelled by role for the KIIs and identified by gender and ward for the FGDs.

### Theme 1: Vaccination patterns and Community acceptability

All key informants reported gradual increasing trends in vaccine acceptance for both children and livestock over the last 5-10 years. The Expanded Programme on Immunization (EPI) nurse reported sustained coverage.

> *“There are times when we are above 85% and at times at 85%…so that is the range between 85-95%” [Subcounty EPI Nurse, KII]*

This was confirmed by the subcounty public health officer who described the improvement as ward specific, confirming an increased uptake in the three study wards:

> *“The community is gradually accepting to vaccinate their children…just a few that may need more sensitization. They are the ones picking up really fast as compared to Matapato North and South.” [Subcounty Public Health Officer, KII]*

For the livestock, the veterinary officer described a meaningful shift from government-led to farmer-led demand:

> *“Earlier most of the time it was mainly centered from the government doing vaccination campaigns. But more and more you’re finding farmers now inquiring about vaccination on their own, privately. I could say the trend has been on an increasing trajectory.” [Subcounty Veterinary Officer, KII]*

Contrary to this sentiment, a farmer from Dalalekutuk ward reported a perceived decline in vaccination campaigns:

> *“Zamani walikuwa wanadunga sana sana wa ng’ombe; sikuhizi ni kama hawadungi…labda wewe mwenyewe uwaite wakudungie ng’ombe wako. (Long ago they used to vaccinate cattle a great deal; nowadays it is as if they do not vaccinate …maybe you yourself have to call them to come vaccinate your cattle.) [Farmer, Dalalekutuk ward, KII]*

### Theme 2: Barriers towards accessing Vaccination services

Transport costs and proximity were the most cited challenges in both FGDs and KIIs. Female FGD participants from Purko and Dalalekutuk wards described poor roads and high motorcycle fares compounded by the long distance to health facilities. The bishop validated this finding.

> *“My opinion is that vaccination needs to be brought nearer to people because in Kajiado the people cover long distances to access vaccination and the issue of transport is a barrier. Vaccines are available but due to distance they are not easily accessed by everyone.” [Bishop, Inter-Religious Council of Kenya, Kajiado county, KII]*

Shortage of Veterinary and Healthcare Worker was documented in both subcounty and household levels. The Officer in charge of vet stores quantified the insufficient staffing:

> *“Sometimes kuna challenge juu hao officers ni few…unapata there are like in Kajiado Central kuna watano” (Sometimes there are challenges because the officers are few… you find there are like five in Kajiado Central) [Officer in charge of vet stores, KII]*

The public health officer also confirmed that the 585 Community health providers across the subcounty were deficient, with even some wards lacking operational community health units. The Sheikh also corroborated similar sentiments citing an extreme of one health worker covering two wards:

> *“In Ildamat we have level two, just one…with one person in. He needs to go maybe to the referral. And all the way from the other ward…Dalalekutuk…an outbreak can spread easily.” [Sheikh, Inter-Religious Council of Kenya, Kajiado county, KII]*

Vaccine supply chain challenges were a systemic barrier at both levels. The EPI nurse mentioned stockouts and fuel funding gaps:

> *“Sometimes we lack fuel to go and collect the vaccine since there is no allocated budget…so sometimes we depend on partners and the sub-county fuel.” [Subcounty EPI Nurse, KII]*

For the livestock, the Veterinary officer identified monopoly in production of vaccines and counterfeit products especially for the expensive FMD vaccine:

> *“Counterfeit vaccines are there…especially on expensive vaccinations like foot and mouth. People get the fake vaccines knowing that they aren’t.” [Subcounty Veterinary Officer, KII]*

The officer in charge of vet stores confirmed an FMD campaign in January had been delayed due to lack of stock:

> *“For now by the way we were supposed to do vaccination in January but we have not yet received the vaccines” [Officer in charge of vet stores, KII]*

Economic barriers were identified at both institutional and community levels. The bishop described a socioeconomic disparity in livestock vaccination access noting that wealthier households called veterinarians to their farms while the poorer ones couldn’t. The farmer directly linked vaccine access to financial capacity.

> *“Inategemea kama ukona pesa” (‘It depends on whether you have money’) [Farmer, Dalalekutuk ward, KII]*

Lack of an adequate notice period was also mentioned in both FGDs and KIIs. Both male and female FGD participants requested advanced (at least one week) notice period before the campaigns. The bishop confirms the current practice is not sufficient:

> *“Sometimes you can even be called early in the morning and you are not well prepared. They need to give notice so that we can prepare the people.” [Bishop, Inter-Religious Council of Kenya, Kajiado county, KII]*

Marginalization of border communities was a KII finding. The EPI nurse identified Meto (Matapato South) as an area where communities live across the Kenya-Tanzania border and has limited access to immunization. Sheikh further elaborated this:

> *“When you go in a place like Meto… it’s in Matapato South. You will find there are some families. They live in Tanzania. Even their school, they go all the way to Tanzania. So…you find they see the Tanzania angle, they see the Kenyan side.” [Sheikh, Inter-Religious Council of Kenya, Kajiado county, KII]*

### Theme 3: Knowledge, Attitudes and Perceptions of Vaccines

Religious leaders provided information on community beliefs and misconceptions. Sheikh identified two narratives within the community including mechanism of transmission

> *“The community was just saying: when you take the vaccine, you will not get a child. And also: these things may make you older before your age. These ideas, they get from among themselves, because it’s a social connection within the community. It’s a narrative.” [Sheikh, Inter-Religious Council of Kenya, Kajiado county, KII]*

The bishop identified the ‘*Kavonokia*’ sect (which rejects conventional medicine and education) as a minor group but locally active and they refuse vaccines due to their religious beliefs.

Gendered decision-making authority was identified as a key mechanism linking household attitudes towards vaccination.

> *“Livestock is everything to the community so they give it a lot of attention. Men believe that livestock belong to them. If it’s the issue of vaccination for children, the women are left to do that work since they believe vaccination belongs to women and children.” [Bishop, Inter-Religious Council of Kenya, Kajiado county, KII]*

Vaccine trust or distrust did not distinguish child or livestock vaccines. Male FGD participants described the outcome of livestock vaccination as proof of concept for child vaccination:

> *“If the cow is vaccinated and stays healthy, I know vaccination works…so when they say vaccinate the child, I believe it also works. It is the same thing – keeping them safe.” [Male FGD, Dalalekutuk]*

Livestock abortion was a concern raised by male FGD participants from Idamat and Purko wards which was confirmed to have pharmacological grounding.

> *“Rift Valley Fever has a slight effect…it causes abortion in some of the animals. And now that is a really negative perception if all the other vaccines are matched with it, because they don’t really know the difference. And if it happens, it’s passed through word of mouth.” [Subcounty Veterinary Officer, KII]*

Vaccine seeking and uptake was primarily motivated by outbreaks. The public health officer described using visible diseases as educational tools:

> *“We have cases of children with polio being used as a case-study to inform those who either refuse or are hesitant. When the community observes a measles case within their area then they will believe that the disease is there and they will plan to take up the vaccine.” [Subcounty Public Health Officer, KII]*

The farmer confirmed proactive preventive measures taken for FMD vaccination based on epidemic occurrence:

> *“Ukiskia inaspread unadunga wanyama kabla hujaona” (‘When you hear it is spreading, you vaccinate before you see it’) [Farmer, Dalalekutuk ward, KII]*

COVID-19 vaccine hesitancy was a distinct topic from routine child immunization hesitancy.

> *“There was a time they declared there is no more COVID. They are not seeing any infections and they say there is no need for them to be vaccinated” [Subcounty EPI Nurse, KII]*

The public health officer corroborated this finding confirming COVID-19 and Measles were key in the context of vaccine hesitancy, whereas under five routine immunizations were administered with low resistance. Furthermore, the Sheikh described COVID-19 fatigue extending into the human papillomavirus (HPV) vaccine resistance among girls.

Trust in government vaccines as opposed to privately sourced products from agrovets was a noteworthy facilitating factor.

> *“Last year agrovets have complained…kwa sababu farmers hawataki agrovets. Wanataka hii ya county, juu wanasema it’s original” (Last year agrovets complained… because farmers do not want agrovets; they want vaccines provided by the county because they say it is original’) [Officer in charge of vet stores, KII]*

### Theme 4: Traditional Remedying

Ethnoveterinary practices were extensively described. Female FGD mentioned ‘*Kubobo’* which involved placing hot ash on the animal’s back to cure East Coast Fever (ECF), washing with urine for Footrot (*Eelei*), using salt and oil for Orf, Powdered sugar for Infectious Bovine Keratoconjunctivitis (IBK) and severing the ears for Trypanosomiasis. Male FGD described ash application for both ECF and Anthrax. The subcounty veterinary officer confirmed that traditional practices were mainly used for treatment and did not act as a barrier towards vaccination.

For children, male participants mentioned specific medicinal plants: *Oleeturot, Empere, Epapa, and Enkii* while female participants listed herbs (Kokula, Olosusiaye, Olkinye, Etulelepwa), prayer and traditional healers as alternatives towards promoting the health in children. The bishop confirmed that herbal medicine was used primarily due to perceived affordability and safety. None of the participants reported traditional remedying as substitutes for vaccination, rather they complemented biomedical care.

### Theme 5: Facilitators and recommendations to increase vaccine uptake

Trusted communication channels were the main facilitator. Female FGD named Radio stations and public address systems that use the Maa language, while the male FGD emphasized on Community Health Volunteers (CHV) and local chiefs citing formal seminars as not being accessible. The EPI nurse elaborated on a proven structure:

> *“We have the community health workers who are helping us in community education. We do health talks every morning before we start our services. We also do community dialogues where we mainly target women and teach them the importance of immunisation.” [Subcounty EPI Nurse, KII]*

Religious leaders are key vaccine champions hence a powerful facilitator. The Sheikh described a ‘co-creating participation’ model where communities are asked their views before programmes are designed, he highlights this as essential towards sustainable acceptance.

> *“We go to the community…we open up a preliminary asking ‘What is your view?’ we call it co-creating participation. We co-create so people say we need this… Religious leaders take vaccines publicly…we start as an example. They see we are being vaccinated, so yes, safe.” [Sheikh, Inter-Religious Council of Kenya, Kajiado county, KII]*

Conversely, the Sheikh also warned that the same religious authority can be used to discourage vaccination:

> *“As a religious leader, when I stand on the podium, and then it’s just two word, ‘shetani ashindwe’ that’s the end of it…so whatever you are bringing for us is shetani…” [Sheikh, Inter-Religious Council of Kenya, Kajiado county, KII]*

Decentralization of service delivery was a structural recommendation that emphasized on extending vaccinations to schools, markets, churches and mosques [Bishop]; increasing the community outreach and home visits [EPI nurse; Female FGD]; Farm visits by veterinarian [Female FGD, Purko ward] and construction of additional crush facilities [Male FGD; Officer in charge of vet stores, KII]

Advanced campaign notice of at least one week was the most actionable recommendation across both KIIs and FGDs. Male FGD requested advanced baraza announcement prior to livestock campaigns so that herds could be gathered from remote grazing areas.

Reforms on supply chain was a key recommendation by the Veterinary Officer in charge of Kajiado central subcounty. She recommended breaking the KEVEVAPI monopoly through licensing additional importers; establishing distributors for the subcounty vaccines so as to reduce counterfeit drugs; investing in thermostable vaccines for solar powered cold chain systems and securing budgets for fuels to facilitate vaccine delivery and collection.

> *“If we could improve the technology of the vaccines to room-temperature stable, it will go a long way because we’ll no longer be using refrigerators, we’re thinking of solar. And we could be having agents so that it’s not only in Kabete… KEVEVAPI should have an agent somewhere like Kajiado town. That could ease the counterfeit products in the market.” [Subcounty Veterinary Officer, KII]*

## Integration and COM-B

Quantitative data (n= 180) were integrated with qualitative data (FGDs n=31 and KIIs n=7) through convergent triangulation. Table 7 summarizes key findings interpreted through the COM-B framework, identifying points of convergence (where both corroborate one another) and divergence (where findings differ).

**Table 7.**
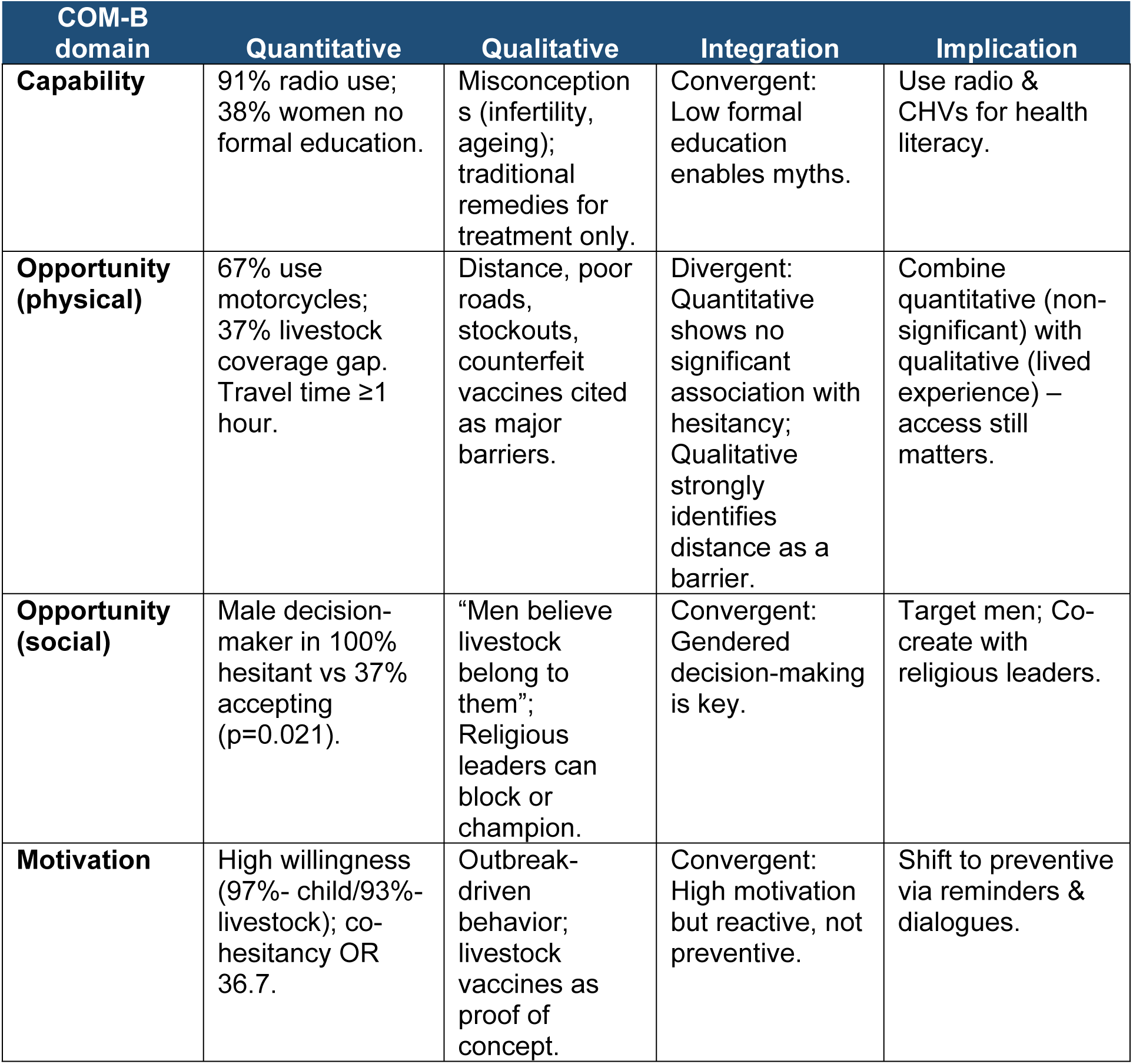
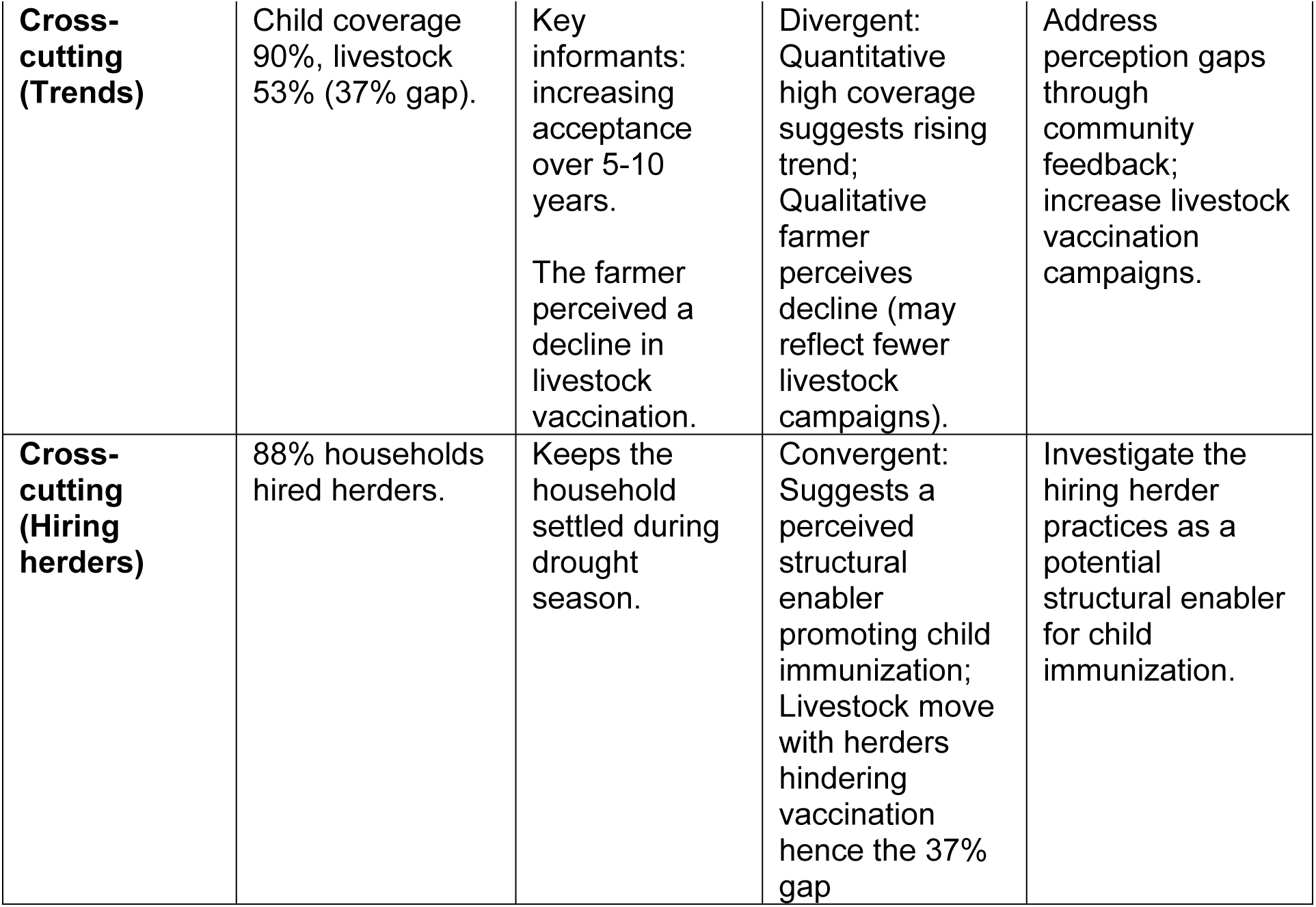
Joint display of integrated findings mapped to the COM–B framework.

## Discussion

This mixed methods study identified factors associated with child and livestock vaccine access among the Maasais in Kajiado central subcounty, Kenya. Our study identified three findings differentiating it from existing literature. First, hiring herders (88% of the households reported) was associated with higher child immunization coverage, suggesting an unintentional structural enabler. Second, the gap between child immunization coverage (90%) and livestock vaccination coverage (53%) despite comparable willingness (97% and 93% respectively) implicates supply chain and systemic failure rather than rejection by the community. Third, vaccine hesitancy co-occurs across both children and livestock within similar households which appear associated with male gendered decision making. However, the sample size was very small and so this finding should be interpreted cautiously.

Previous studies among the Maasai community in Kajiado have documented consistently low child immunization coverage, attributed primarily to household mobility, cultural norms, and constrained access to services(7,9). Contrary to this, our study observed an 90% child immunization coverage in Kajiado Central, notably higher than prior estimates therefore challenging the assumption that mobility remains among the dominant barriers in this specific context. The high prevalence of households hiring herders during the cool-dry season (88%) may signal a broader livelihood transition toward sedentarization that incidentally aligns these communities more closely with conventional facility-based immunization design. We hypothesize this structural shift may partially explain the higher-than-expected coverage, though prospective research is needed to establish whether hiring herders could function as a scalable, community-embedded enabler of vaccine access in pastoral settings.

However, hiring herders may also inadvertently reduce livestock vaccine access. When they take animals to distant grazing grounds (far from established vaccination crushes) the logistical challenge of gathering herds increases, and maintaining the cold chain over long distances becomes substantially more difficult(2). Therefore, addressing the livestock vaccination gap may require vaccination strategies that account for mobility such as vaccinating at seasonal watering points(16).

Despite high willingness (>90%) to vaccinate both livestock and children, livestock vaccination coverage was 37% lower than child immunization, pointing towards systemic failures. Qualitative data from this study indicated that vaccination in Kajiado Central Subcounty was predominantly reactive, triggered by outbreaks. For instance, healthcare workers actively used measles and polio cases as educational tools, while farmers initiated FMD vaccination campaigns upon epidemic occurrence, likely due to resource constraints. This pattern, where high willingness to vaccinate co-exists with low routine coverage, is consistent with a scoping review which found that joint human-animal vaccination campaigns were often designed around polio or outbreak response(2). In addition, key informants from the veterinary department described systemic failures including: KEVEVAPI’s vaccine production monopoly, counterfeit FMD vaccines in the market, lack of resources for vaccine distribution at subcounty and ward level(17). Crucially, at the time of the interviews, a planned FMD vaccination campaign had been postponed due to lack of stock. This stands in stark contrast to child immunization, which benefits from a more established infrastructure of community health promoters, routine outreach programs, and a functional cold chain system. Similar supply-chain constraints have been documented in pastoral livestock vaccination systems across East Africa(2,6) and a scoping review on joint human-animal vaccination strategies in Africa confirmed that animal health outcomes are consistently overlooked and supply challenges remain unaddressed(2). These findings collectively reinforce the need for livestock supply-chain investment: breaking the KEVEVAPI’s monopoly, strengthening Veterinary Medicine Directorate (VMD) regulatory framework, investing in thermostable vaccines for solar-compatible cold chain systems, and securing dedicated budgets for vaccine transport in remote areas.

The study found potential signals of co-occurrence of child and livestock vaccine hesitancy within the same household points to a shared social mechanism rather than independent attitudinal patterns. While qualitative data described child vaccination as the responsibility of women and livestock vaccination as men’s domain, quantitative data revealed that in all child–hesitant households, the male partner was the ultimate decision–maker. These seemingly divergent findings can be reconciled: women manage routine child vaccination, but their decisions are accepted as long as they align with male authority. This pattern explains why child and livestock hesitancy cluster in the same households: the same male authority governs both domains ultimately. Similar gendered patterns where men control vaccination decisions have been described among Ethiopian pastoralists(18). Notably, male FGD participants described healthy vaccinated livestock as proof of concept for child immunization, meaning that positive experiences in one domain can spill over into the other, and conversely that negative experiences or distrust can do the same. A scoping review noted that joint vaccination campaigns have consistently treated human and animal health as separate entities, focusing predominantly on child immunization while giving little attention to livestock vaccination(2). This study supports a One Health approach that targets male decision-makers directly and simultaneously across both domains, rather than through parallel and disconnected programmes.

Religious leaders represent a double-edged social opportunity that warrants deliberate programmatic engagement. The Sheikh informant described how a single pulpit declaration, *“Shetani Ashindwe”* (Satan be defeated), can instantly reframe vaccines as evil and collapse community trust. The same authority, however, can be channeled constructively through what the Sheikh described as a co-creation model: pre-campaign community consultation, public vaccination of religious leaders as visible proof of safety, and community co-design of health messaging. This model directly addresses demand-side challenges documented in Chad, where lack of dialogue and culturally distant health staff were primary drivers of hesitancy(5). Its replicability is supported by evidence from Kano State, Nigeria, where a comparable approach persuaded over 80% (n=7927) of households that had initially rejected vaccines, and from Angola, where religious leaders and traditional healers were central to public health communication(19). Engaging the religious leaders in Kajiado Central, as a formal partner in vaccination campaign design offers an actionable pathway grounded in existing community infrastructure(20).

Despite high community acceptance, misconceptions persisted that warrant targeted communication responses. Vaccines causing infertility and premature ageing were cited as community narratives, consistent with findings from mobile pastoralists in Chad where similar misconceptions drove hesitancy(5). In the livestock domain, concerns about abortion following Rift Valley Fever vaccination were overgeneralized to all livestock vaccines, creating broader hesitancy. These concerns have pharmacological grounding: older live-attenuated vaccines such as the Smithburn strain posed a documented risk of abortion and teratogenic effects in pregnant ewes(21). However, safer alternatives such as the RVF Clone 13 vaccine have since been developed and validated for use in pregnant animals(22). Addressing this requires animal health practitioners and trusted community messengers to actively distinguish between old and newer vaccine types, deliver corrective messaging through household visits, and provide live demonstrations where feasible. Radio communication in *Maa* language is already the dominant channel for child immunization information among 91% of participants and word-of-mouth through village elders and churches (71% for livestock) represent the most efficient existing channels through which corrective messages can reach these communities.

Beyond knowledge and attitudes, physical access to vaccination services remained a structural barrier. Two thirds of participants used motorcycles to reach health facilities at a median cost of Ksh 150 (US$1.2), while 47% walked, reflecting both the terrain and the absence of closer alternatives. Among child-hesitant households, 67% required over one hour to reach a facility. This mirrors findings from Garissa (Lagdera) where 79% of nomadic mothers cited distance as the primary barrier to vaccination(23). Decentralizing service delivery to schools, markets, churches, and mosques, complemented by increased community outreach, home visits, and farm visits by animal health service providers emerged as the most consistently recommended structural solution across both FGDs and KIIs. For livestock campaigns specifically, advance baraza announcements of at least one week were requested by male FGD participants to allow herds to be gathered from remote grazing areas, a simple, low-cost operational adjustment with potentially significant coverage implications.

Ethnomedical and ethnoveterinary practices were extensively described but used exclusively for treatment, and were reported as complementary to rather than competitive with vaccination. This challenges common assumptions that traditional medicine undermines immunization uptake(5,13), and is consistent with findings from Oloitoktok Subcounty, Kajiado, where traditional practices similarly did not act as barriers to livestock vaccination(24). This finding suggests that traditional remedying should not be treated as obstacles, but rather as parallel practices that coexist with biomedical care in this context.

COVID-19 vaccine hesitancy appeared to be distinct from routine under-five immunization hesitancy, which experienced minimal disruption. This aligns with evidence from Rwanda, where strong pro-vaccination social norms buffered routine child immunization from pandemic-related disruption(25). However, COVID-19 fatigue did spill over into HPV vaccine resistance among adolescent girls in Kajiado Central, a pattern consistent with global evidence that the pandemic reduced HPV vaccination uptake (26,27) warranting targeted adolescent health communication strategies that decouple HPV vaccination from COVID-19 narratives.

This study had several limitations. As a cross-sectional study design, it does not permit assessment of temporality or causal inference. Additionally, reliance on self-reporting for livestock vaccination may introduce social desirability bias, although 91% of participants presented child immunization cards, corroborating self-reported child vaccination data. The snowball sampling technique used may introduce selective bias and limit generalizability beyond the study wards. The small number of hesitant households in both child (n=6) and livestock (n=13) outcomes were insufficient to support multivariable analysis, meaning all hesitancy-related findings are exploratory and should be interpreted with caution. Despite these limitations, the mixed-methods design with convergent triangulation strengthens internal validity and enhances transferability to comparable pastoralist contexts in East Africa.

This study observed child immunization coverage considerably higher than previously documented among Maasai pastoralists in Kajiado, which alongside the high prevalence of hiring herders, raises a hypothesis of broader sedentarization that warrants further investigation as a potential structural enabler of vaccine access in pastoral settings.

Quantitative data showed consistently low livestock coverage despite high willingness, triangulated with qualitative accounts of production monopoly, counterfeit vaccines, stockouts, understaffing, and absent ward-level distribution infrastructure, collectively rule out community rejection as the primary explanation. Vaccine hesitancy, though low, appeared to present across children and livestock within the same households, pointing to a shared household-level dynamic mediated by male decision-making authority that a siloed programmatic approach would not address.

Therefore, we recommend a coordinated One Health response operating on two tracks. Supply-side reforms which include breaking the KEVEVAPI monopoly, licensing additional importers, strengthening anti-counterfeit regulation, investing in thermostable vaccines for solar-compatible cold chain systems, and securing dedicated budgets for vaccine transport up to the ward level. Further research is needed to further investigate the co-occurrence of child and livestock vaccine hesitancy. If our findings hold true, then demand-side efforts must deliberately engage male decision-makers as the common gateway to both child and livestock vaccination. Such efforts must leverage the co-creation model championed by religious leaders already active in Kajiado Central subcounty, decentralize service delivery to markets, schools, churches, and mosques, and provide at least one week of advance campaign notice. Collectively, these measures have the potential to offer contextually grounded and transferable pathways toward equitable vaccination coverage in pastoral communities across East Africa.

## Data Availability

The minimal data set [and accompanying code, where generated] is available at (https://drive.google.com/drive/folders/1N1ujg9eRnYpLOtOGWMbQyPS1CTuTbvEB?usp=drive_link)

## Acknowledgement

The authors thank the County Government of Kajiado for granting administrative permission. They are grateful to the Kajiado Central Subcounty public health and veterinary officers for their support and to all study participants for their time and insights.

